# Associations between social support and verbal memory: 14-year follow-up of the English Longitudinal Study of Ageing cohort

**DOI:** 10.1101/2020.07.02.20145037

**Authors:** Shaun Scholes, Jing Liao

## Abstract

**Objective:** Examine the longitudinal associations between social support and verbal memory among community-dwelling adults.

**Participants:** In the English Longitudinal Study of Ageing, 10,837 participants aged 50-89 years were assessed at wave 1 (baseline: 2002-03) and followed-up over 14-years to wave 8 (2016-17).

**Methods:** Verbal memory was assessed at each wave by tests of immediate and delayed word-recall. Positive and negative social support were measured across four types of relationship (spouse/partner, children, friends, extended family). Linear mixed effects modelling examined the between-person (PM) and within-person (PM) associations between social support and verbal memory. Interaction terms were fitted to estimate differences in the rate of change in verbal memory by social support.

**Results:** When we summed scores across all relationships (global social support), higher-than-average (PM) positive social support was associated with higher baseline verbal memory (β_PM_ = 0.015; 95% Confidence Interval (CI): 0.008, 0.025; *p* < 0.001) and with slower decline (β_PM-by-time_ = 0.003; 95% CI: 0.001, 0.005; *p* = 0.003). Longitudinal associations were gender- and source-specific. Among men, slower decline in verbal memory was associated with higher positive social support from children (β_PM-by-time_ = 0.009; 95% CI: 0.002, 0.016; *p* = 0.011) and with lower negative social support from spouse/partner (β_PM-by- time_ = −0.016; 95% CI: −0.031, 0.000; *p* = 0.045) and from friends (β_PM-by-time_ = −0.021; 95% CI: −0.039, −0.004; *p* = 0.018). Among women, higher positive social support from extended family was associated with slower decline in verbal memory (β_PM-by-time_ = 0.008; 95% CI: 0.000, 0.015; *p* = 0.049).

**Conclusions:** Between-person differences in social support were modestly associated with differential decline in verbal memory. Our findings can inform future research studies and intervention strategies designed to maximise the potential role of supportive relationships in promoting healthy cognitive ageing.

## Introduction

Prospective cohort studies have linked cognitive ageing with higher risk of morbidity and mortality (Sabia *et al*., 2010; Batty, Deary and Zaninotto, 2016). Beyond the influences of ageing and depressive symptomology, poor social relationships have been longitudinally associated with higher risk of cognitive decline (Kuiper *et al*., 2016; Kelly *et al*., 2017). Regarding functional aspects of social relationships, provision of negative (but not positive) social support from close ties predicted accelerated cognitive decline in the Whitehall II UK cohort (Liao *et al*., 2014). In the US Health and Retirement Study (HRS), structural aspects of social relationships - being married/partnered and reporting more contact with friends - were independently associated with slower decline in episodic memory (Zahodne *et al*., 2019).

Few studies to date have examined long-term longitudinal associations between social support (positive and negative) and cognitive ageing, and whether these associations are gender and source-specific (Zahodne *et al*., 2019). In an earlier investigation with 8-year follow-up in the English Longitudinal Study of Ageing (ELSA), we showed that positive support from a spouse/partner predicted slower but modest decline in executive function among men but not among women (Liao and Scholes, 2017), indicating that men and women may perceive and utilise social support in different ways (Antonucci and Akiyama, 1987). Associations between poor social relationships and increased risk of cognitive decline were found in a recent meta-analysis to be stronger in studies with 8+ years follow-up (Kuiper *et al*., 2016). Longer follow-up increases statistical power, provides more robust measurement of within-person change, and reduces potential for reverse causality with respect to variables assessed at baseline (Kuiper *et al*., 2016). Using 14-years follow-up in the ELSA cohort (2002-03 to 2016-17), the primary aim of this study is to explore the longitudinal associations between social support and cognitive function (verbal memory). We hypothesise that: (1) higher positive and lower negative social support associate independently with higher baseline verbal memory and with slower decline; (2) both between-person differences and within-person change in social support independently predict baseline levels and change in verbal memory; and (3) associations differ by gender and by source of social support.

## Methods

### Study design and participants

ELSA is an ongoing study of adults living in private households in England; 11,391 sample members (born before March 1, 1952) participated in wave 1 (2002–03; 67% response rate). A detailed description of the goals, design and methods of ELSA is available elsewhere (Steptoe *et al*., 2013). Data collection takes place every two years via face-to-face interviews and self-completion questionnaires. To minimise potential for reverse causality (Zaninotto *et al*., 2018), we excluded participants at wave 1 with doctor-diagnosed Alzheimer or Parkinson disease, dementia, or serious memory impairment. Proxy respondents, those aged 90+, and those with missing baseline cognition data were also excluded.

### Consent and ethical approval

ELSA participants provided signed consent for taking part in the study and for linkage to mortality data; ethical approval was granted by the London Multicentre Research Ethics Committee (MREC/01/2/91).

### Assessment of verbal memory via word-recall tests

Verbal memory was our chosen outcome measure as tests were administered at each wave. Participants were presented with a list of 10 words that were read out by a computer at the rate of 1 word for every 2 seconds (Steel *et al*., 2002). Participants were then asked to recall as many words as they could (immediate recall); they were asked to recall these words after an interval during which they completed other cognitive tests (delayed recall). The between-test Pearson correlation over the 8 waves ranged from 0.70 to 0.77. Both scores were summed to obtain a composite continuous measure of correctly recalled words (range, 0-20 words); these were normally distributed, suggesting the absence of floor or ceiling effects. Word-recall tests have shown good construct validity and consistency (Baars *et al*., 2009).

### Assessment of social support

Questions on social support were administered at each wave and covered four sources: (1) spouse/partner; (2) children; (3) friends; and (4) extended family. Three questions addressed positive social support: (a) how much they understand the way you feel about things; (b) how much they can be relied on if you have a serious problem; and (c) how much you can open up to them to talk about worries. Responses ranged from “not-at-all” (scored 0) to “a lot” (3). Scores were summed for each relationship (range, 0–9) and summed into an overall ‘global’ score (range, 0–36). Three questions addressed negative social support: (a) how much they criticize you; (b) how much they let you down when you are counting on them; and (c) how much they get on your nerves. Responses were scored as described for positive support (i.e. higher values indicated more negative perceptions). Participants without the relevant social ties were scored zero.

### Confounders

Based on previous research (Zaninotto *et al*., 2018; Yin *et al*., 2019), we identified the following confounders (assessed at wave 1): demographic (age, range: 50-89 years); markers of socioeconomic position (SEP: wealth and education); healthy lifestyle behaviours (smoking, alcohol and physical activity); social participation; physical functioning; and depressive symptoms. Total wealth represented the sum of financial, physical and housing wealth, minus debts, and was grouped into quintiles (lowest to highest). Education status was categorised as low (compulsory schooling); medium (up to high school) and high (university degree or higher). Smoking status was classed as current cigarette smoker or not; frequency of alcohol consumption in the past year was classed as less than daily or daily. Participants were asked how often they engaged in moderate and in vigorous sports/activities: we classed participants as physically active or inactive. A binary social participation score was created based on involvement in a list of activities related to civic participation, leisure activities and cultural engagement. Participants were classed as having a mobility limitation if they reported having difficulties in performing any of six basic activities of daily living tasks (ADL) (Katz *et al*., 1963). For depression we used a binary indicator based on having a score of 4+ depressive symptoms on the 8-item Center for Epidemiologic Studies Depression Scale (CESD-8)(Radloff, 1977).

### Statistical analyses

#### Missing data

To evaluate item nonresponse bias, the distributions of key variables were examined according to whether participants had missing baseline data on positive social support. Distributions were also examined according to whether participants completed 1, 2-5, and 6-8 assessments of verbal memory during follow-up to examine potential attrition bias. Differences in distributions were evaluated using Wald (continuous variables) and Rao-Scott chi-square tests (categorical variables).

#### Descriptive statistics

Means and standard deviations (SD) for continuous variables and percentages for categorical variables were used to present the characteristics of the analytic sample by study wave.

#### Linear mixed effects modelling

Linear mixed effects (LME) modelling with time-since-baseline as timescale (range, 0-7) was used to estimate baseline levels and change in verbal memory (Laird and Ware, 1982). LME models use all available data, account for intraindividual correlation, and is robust under the assumption that data is missing at random (MAR). Random intercepts and slopes were included (via random effects) to account for unexplained variation between participants in baseline verbal memory and in its rate of change, respectively; an additional term estimated intercept-by-slope covariance. Analyses were weighted using the wave 1 weight to ensure that the sample was broadly representative of the community-dwelling English population aged 50+ years at baseline. We conducted three model-based analyses.

##### 1. Unadjusted model

First, we estimated the baseline levels and two-year change in verbal memory using time and time-squared (linear and quadratic change, i.e. time^2^); baseline age (centred at 65 years) and age^2^; and their statistical interaction (time × age; time × age^2^) as fixed effects. Including time^2^ allows for non-linear decline in verbal memory; the main effect of age^2^ allows for a steeper decrease in baseline verbal memory with increasing age.

##### 2. Fully-adjusted model: global social support

Second, we added the global social support variables as fixed effects as time-varying predictors. As in our previous study (Liao and Scholes, 2017), we explicitly accounted for stable (‘*trait*’) differences between-persons in their average score and for within-person change (‘*state*’) (Hoffman and Stawski, 2009). Between-person associations were assessed using each participant’s average score during follow-up, centred at the grand mean (hereafter referred to as the person-mean (PM) variable). Within-person (WP) associations were assessed by subtracting each participant’s wave-specific score from their own average (hereafter referred to as the WP variable). PM coefficients represent the change in verbal memory per unit change between-persons in their average level of social support; WP coefficients represent the change in verbal memory per unit change in each participant’s *usual* level of social support (Hoffman and Stawski, 2009).

Separately for positive- and negative-support, the models contained four terms of primary interest: the main effects (PM; WP) and their interaction with time (linear change only); the cross-level interaction terms PM × WP were statistically insignificant (*p* > 0.32) and so were not retained. Evidence of effect modification by gender was weak (*p* = 0.583), and so data for men and women were pooled and our modelling adjusted for gender.

The fully-adjusted model contained terms for time, time^2^, age, age^2^, the positive and negative social support variables (PM, WP), and the potential confounders listed above. Interaction terms for each were included to represent varying rates of change in verbal memory. We adjusted for the number of prior word-recall tests (range, 0-7) to correct for practice effects (Vivot *et al*., 2016); this term also proxies characteristics that influence attrition, which in turn, strongly associate with cognitive task performance (Karlamangla *et al*., 2009, 2017).

To minimise the impact of item nonresponse on statistical precision and power (Sterne *et al*., 2009), we used the multiple imputation using chained equations (MICE) method (Rawlings *et al*., 2017). Conditional imputation (Royston and White, 2011) was used to impute social support scores conditional on the relevant social ties being present. The imputation models used as predictors all variables in our primary analysis (including baseline verbal memory) plus auxiliary variables also assessed at baseline. LME model estimates were combined across 10 imputed datasets using Rubin’s rules.

##### 3. Fully-adjusted model: gender- and source-specific analysis

In our third analysis, we decided a-priori to fit gender and source-specific models.

### Sensitivity analysis

Two sensitivity analyses assessed the impact of biases associated with attrition on our main findings. First, we calculated inverse-probability weights (IPWs) based on study dropout (conditional on survival), then used these weights to model the outcome using a generalized estimating equation (IPW-GEE) linear regression model with an independent working correlation structure and robust variance estimation (Daza, Hudgens and Herring, 2017). Secondly, we reran the LME models on the subset of participants who completed all 8 waves (‘completers’), using the weight provided with the publicly available datasets that adjusts for cumulative attrition since wave 1.

Data was analysed using Stata v15.1 (Stata Corp LP, College Station, Texas). Statistical significance tests were based on two-sided probability (*p* < 0.05); no adjustment was made for multiple comparisons.

## Results

### Analytic sample

Flowcharts showing derivation of the analytic samples are provided in Figures S1-S2. The main analytic sample comprised 10,837 participants aged 50-89 years. On average, these had 4.8 (SD 2.7) assessments of verbal memory during follow-up; 1847 (17%) took part only at wave 1; 3282 (30%) completed all 8 waves.

### Missing data due to item non-response

Baseline data on positive social support was missing for 780 (7.2%) participants (Table S1). Higher levels of missing data were associated with older age; lower scores of verbal memory; low SEP; cigarette smoking; physical inactivity; having 1+ mobility limitations, and having 4+ depressive symptoms (all *p* < 0.001).

### Missing data due to unit nonresponse

Completing 6-8 assessments of verbal memory during follow-up was associated with younger age; better verbal memory performance at baseline; high SEP; healthy lifestyles; having no mobility limitations and being non-depressed (all *p* < 0.001; Table S2).

### Descriptive statistics

Table 1 shows the characteristics of the key variables for those remaining in the study during follow-up. Mean age at wave 1 was 64.8 years, fewer than half were male (45%), and over one-third had completed no more than compulsory schooling (42%). Over time, age decreased on average by 4.5 years, whilst verbal memory performance increased on average between waves 1 and 2 but remained at a similar level thereafter.

**Table 1.** Analytical sample characteristics by study wave

| **Characteristics** | **Wave 1** | **Wave 2** | **Wave 3** | **Wave 4** | **Wave 5** | **Wave 6** | **Wave 7** | **Wave 8** |
| --- | --- | --- | --- | --- | --- | --- | --- | --- |
|  | 2002-03 | 2004-05 | 2006-07 | 2008-09 | 2010-11 | 2012-13 | 2014-15 | 2016-17 |
|  | Mean (SD) | Mean (SD) | Mean (SD) | Mean (SD) | Mean (SD) | Mean (SD) | Mean (SD) | Mean (SD) |
| Participants (n) | 10,837 | 8423 | 7195 | 6257 | 5831 | 5293 | 4573 | 3929 |
| **Time-varying** |  |  |  |  |  |  |  |  |
| Word recall <sup>a</sup> | 9.4 (3.5) | 9.9 (3.6) | 10.0 (3.7) | 10.0 (3.7) | 10.0 (3.8) | 10.2 (3.8) | 9.9 (3.8) | 9.9 (3.9) |
| Positive support <sup>b</sup> | 22.4 (7.5) | 22.6 (7.5) | 23.0 (7.4) | 22.3 (7.4) | 21.9 (7.4) | 22.0 (7.4) | 22.6 (7.4) | 22.0 (7.4) |
| Negative support <sup>b</sup> | 6.5 (4.8) | 6.6 (4.8) | 6.5 (4.8) | 6.4 (4.8) | 6.4 (4.6) | 6.1 (4.5) | 6.1 (4.7) | 6.0 (4.6) |
| **Time-invariant** |  |  |  |  |  |  |  |  |
| Age, years | 64.8 (9.9) | 64.3 (9.6) | 63.8 (9.4) | 63.1 (8.9) | 62.4 (8.6) | 61.9 (8.2) | 61.1 (7.8) | 60.3 (7.3) |
| Male (%) | 45 | 45 | 45 | 44 | 44 | 44 | 43 | 43 |
| Low level of education (%) | 42 | 39 | 37 | 35 | 34 | 33 | 30 | 29 |
| Lowest wealth (%) | 19 | 18 | 17 | 16 | 16 | 16 | 15 | 15 |
| Current smoker (%) | 18 | 17 | 17 | 16 | 16 | 16 | 16 | 15 |
| Daily alcohol drinker (%) | 28 | 29 | 29 | 29 | 29 | 30 | 30 | 30 |
| Physically inactive (%) | 18 | 16 | 14 | 13 | 12 | 11 | 10 | 9 |
| No social participation (%) | 30 | 28 | 27 | 26 | 25 | 24 | 23 | 23 |
| CESD-8: 4+ (%) | 16 | 15 | 15 | 14 | 14 | 13 | 13 | 12 |
| ADL: 1+ (%) | 20 | 18 | 18 | 16 | 16 | 15 | 14 | 13 |

Abbreviations: ADL = activities of daily living; CESD-8 = 8-item Centre for Epidemiological Studies Depression Scale; SD = standard deviation.
<sup>a</sup> Sum of correctly recalled words (immediate and delayed).
<sup>b</sup> Global social support (scores summed from all 4 sources: spouse/partner; children; friends; extended family). Estimates are unweighted; the number of participants (n) is shown for those with non-missing verbal memory scores: participants with missing data on the other variables are excluded from this table.

### Unadjusted model

The unadjusted LME model showed non-linear accelerated decline in verbal memory (β_time_ ^2^: *p* < 0.001) and faster decline at older ages (*p* for all terms < 0.001) (Table S3). On average, verbal memory performance decreased by 1.6 (95% CI: −1.7, −1.5; *p* < 0.001) during the 14-year follow-up.

### Global social support and verbal memory

Adjusting for global social support, better verbal memory performance at baseline was associated with being younger; female; having a degree or higher qualification; higher wealth; being non-depressed; daily alcohol consumption; being physically active; and being socially active. Faster decline in verbal memory was associated with increasing age (age and age^2^: *p* <0.001); being male (*p* < 0.001); and cigarette smoking (*p* < 0.001) (Table S4). Separately for positive- and negative-support, Table 2 shows the fully-adjusted coefficients for the main effects of the between- and within-person associations (PM and WP, respectively) and their interaction with time.

**Table 2.** Coefficients from linear mixed effects models of verbal memory by positive and negative social support from all sources

|  | Positive social support |  |  | Negative social support |  |  |
| --- | --- | --- | --- | --- | --- | --- |
| | Estimate<br>($\beta$) | 95% CI | P-value | Estimate<br>($\beta$) | 95% CI | P-value |
| **Fixed effects:** |  |  |  |  |  |  |
| *Between-person:* |  |  |  |  |  |  |
| PM | 0.016 | 0.008, 0.025 | **<0.001** | -0.028 | -0.046, -0.011 | **0.002** |
| PM × time | 0.003 | 0.001, 0.005 | **0.003** | 0.000 | -0.004, 0.004 | 0.985 |
| *Within-person:* |  |  |  |  |  |  |
| WP | 0.001 | -0.009, 0.011 | 0.888 | 0.009 | -0.005, 0.024 | 0.210 |
| WP × time | -0.001 | -0.004, 0.002 | 0.709 | -0.004 | -0.008, 0.000 | **0.046** |
| **Random effects:** |  |  |  |  |  |  |
| Intercept variance | 4.05 | 3.86, 4.25 |  |  |  |  |
| Slope variance | 0.08 | 0.07, 0.09 |  |  |  |  |
| Intercept-by-slope<br>(correlation) | -0.07 | -0.14, -0.01 |  |  |  |  |

Abbreviations: CI = confidence interval; PM = person-mean (between-person associations); WP = within-person. Results in bold indicate $P < 0.05$ .

With regards to stable (‘*trait*’) differences between-persons, higher positive social support was associated with higher baseline verbal memory (β_PM_ = 0.016; 95% CI: 0.008, 0.025; *p* < 0.001) and with slower decline (β_PM×time_ = 0.003; 95% CI: 0.001, 0.005; *p* = 0.003) (Figure 1). Lower negative social support was associated with higher baseline verbal memory (β_PM_ = −0.028; 95% CI: −0.046, −0.011; *p* = 0.002) but not with decline (β_PM×time_: *p* = 0.985). With regards to within-person change (‘*state*’), higher-than-usual levels of negative social support were modestly associated with faster decline (β_WP×time_ = −0.004; 95% CI: −0.008, 0.000; *p* = 0.046).

**Figure 1.**
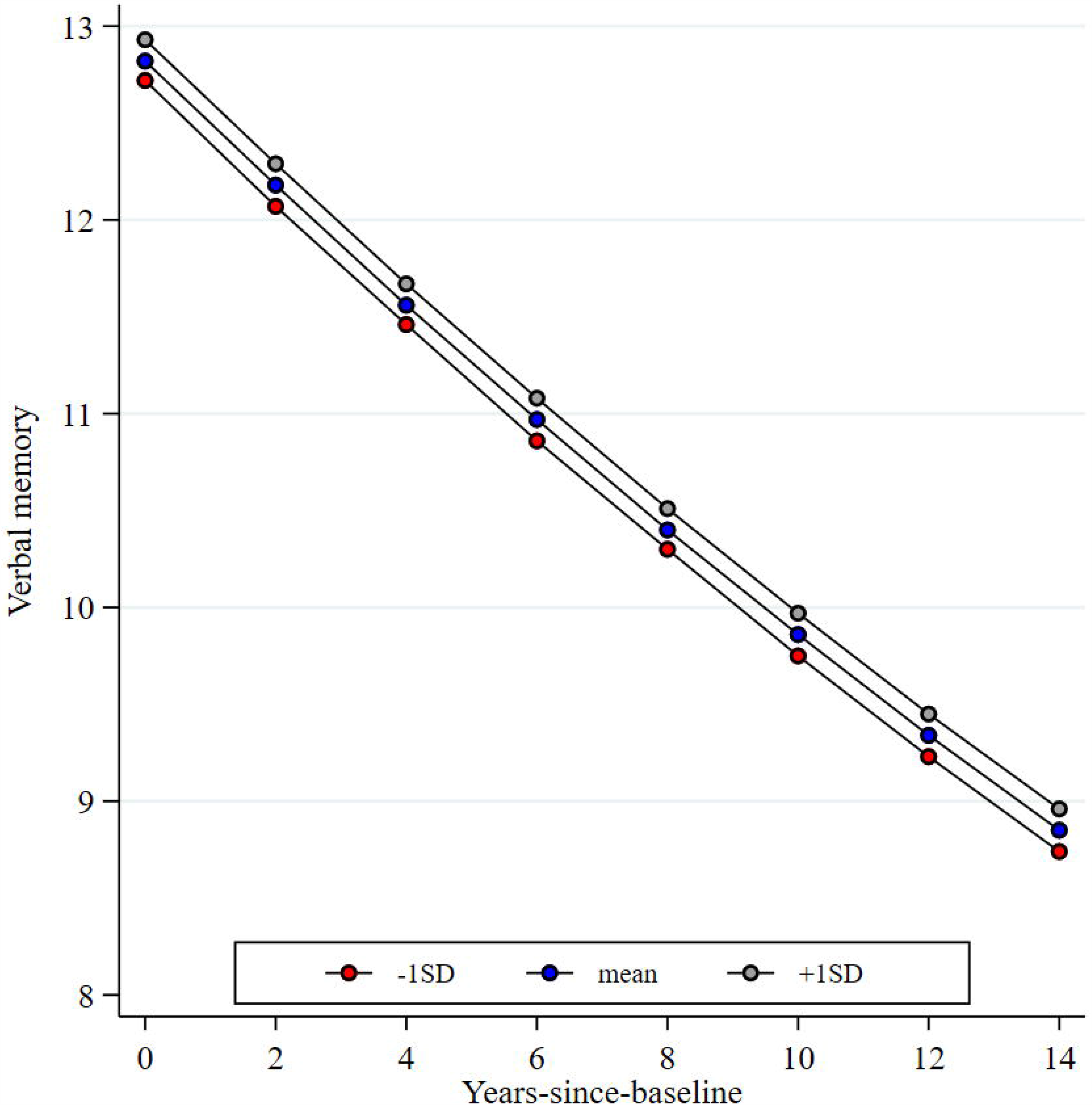
Verbal memory trajectories according to differences between-persons in positive social support. Predicted trajectories in verbal memory as a function of time-since-baseline (wave 1 to wave 8; 0–14 years). Figure shows the predicted trajectory (at the mean and ±1 SD) for levels of positive social support (PM: between-person differences). Slower decline in verbal memory was modestly associated with higher (‘stable’) levels of positive social support (β_PM×time_ *P* = 0.003) (Table 2; Table S4). Trajectories estimated with other variables held constant at reference values (number of prior assessments = 2.7; age at wave 1 = 65; male; university degree or higher; highest wealth quintile; non-smoker; non-daily alcohol consumption; physically active; socially active; no mobility limitations; non-depressed).

### Trajectories in verbal memory by gender and source of support

Among men, slower decline in verbal memory was associated with lower negative social support from a spouse/partner (β_PM×time_ = −0.016; 95% CI: −0.031, 0.000; *p* = 0.045) and with higher positive social support from children (β_PM×time_ = 0.009; 95% CI: 0.002, 0.016; *p* = 0.011) (Figure 2; Table S5). Higher positive social support from friends was associated with higher baseline memory (β_PM_ = 0.054; 95% CI: 0.008, 0.100; *p* = 0.023); lower negative social support from friends was associated with higher baseline levels (β_PM_ = −0.123; 95% CI: −0.209, −0.037; *p* = 0.006) and with slower decline (β_PM×time_ = −0.021; 95% CI: −0.039, − 0.004; *p* = 0.018). Lower negative social support from extended family was associated with higher baseline memory (β_PM_ = −0.128; 95% CI: −0.199, −0.058; *p* = 0.001).

**Figure 2.**
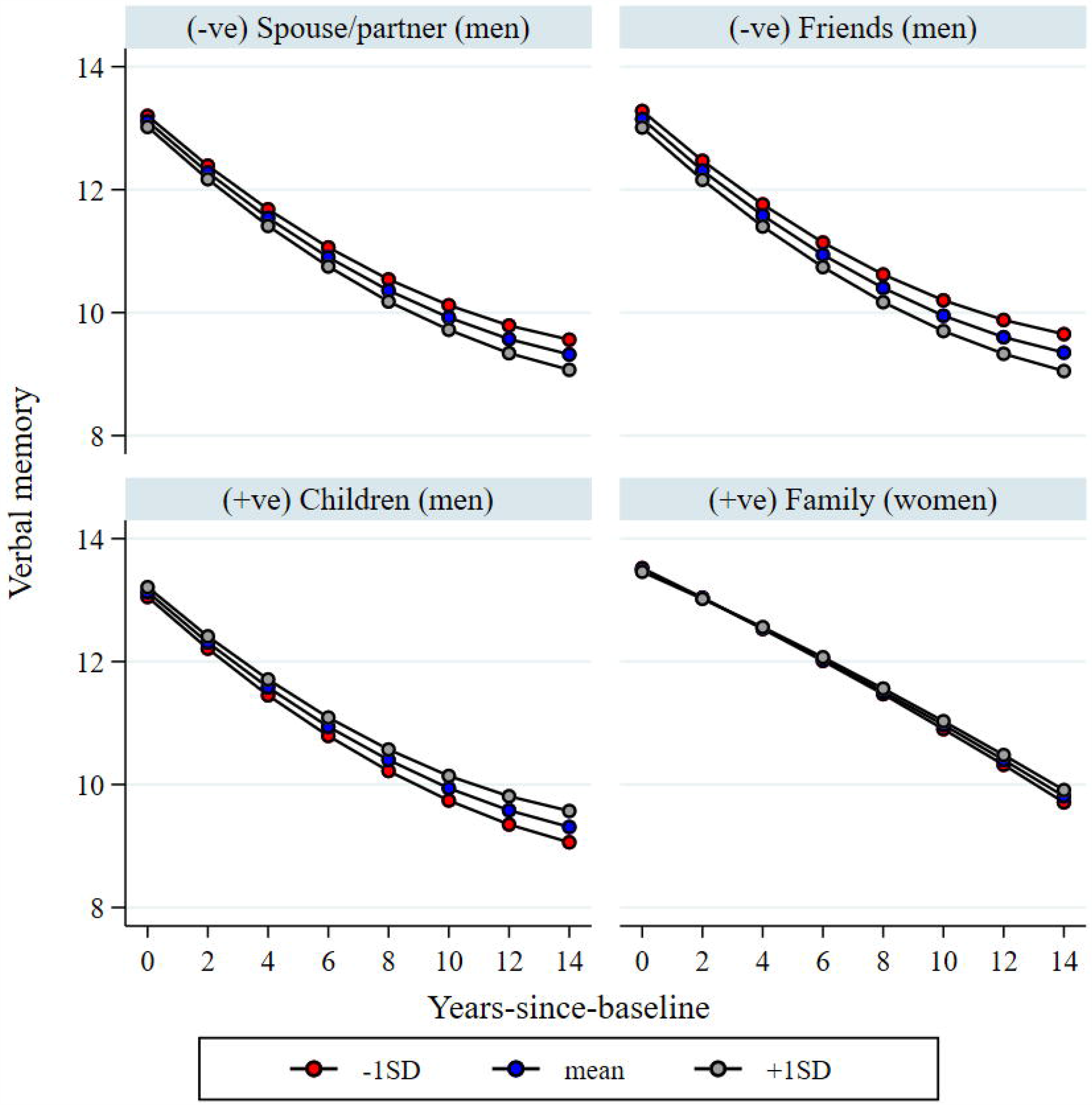
Verbal memory trajectories according to gender and source of social support. Predicted trajectories in verbal memory performance as a function of time-since-baseline (wave 1 to wave 8; 0–14 years). Figure shows the predicted trajectory (at the mean and ±1 SD) for levels of positive (+ve) social support and levels of (-ve) negative social support (PM: between-person differences). Trajectories estimated with other variables held constant at reference values (number of prior assessments = 2.7; age at wave 1 = 65; male; university degree or higher; highest wealth quintile; non-smoker; non-daily alcohol consumption; physically active; socially active; no mobility limitations; non-depressed).

Among women, lower negative social support from children was associated with higher baseline memory (β_PM_ = −0.090; 95% CI: −0.157, −0.023; *p* = 0.009) but not with decline (β_PM×time_: *p* = 0.692). Higher positive social support from friends was associated with higher baseline levels (β_PM_ = 0.104; 95% CI: 0.062, 0.146; *p* < 0.001) but not with decline (β_PM×time_: *p* = 0.559). Higher positive social support from extended family was modestly associated with lower decline (β_PM×time_ = 0.008; 95% CI: 0.000, 0.015; *p* = 0.049).

### Sensitivity analyses

In our first sensitivity analysis, estimates from the IPW-GEE linear regression model showed that an 1-unit increase in the usual level of negative social support was associated with higher baseline verbal memory (β_WP_ = 0.028; 95% CI: 0.009, 0.046; *p* = 0.004), and with faster decline (β_WP×time_ = −0.009; 95% CI: −0.015, −0.002; *p* = 0.009) (Table S6). The corresponding LME estimates were similar in sign but weaker in magnitude (Figure S4).

Secondly, estimates from LME models using data only from those who completed all 8 waves were less precise, but showed similar findings to our main analysis. Slower decline in verbal memory was associated with higher positive social support (β_PM×time_ = 0.004; 95% CI: 0.002, 0.007; *p* < 0.001) whilst higher-than-usual levels of negative social support were modestly associated with faster decline (β_WP×time_ = −0.005; 95% CI: −0.011, 0.000; *p* = 0.049) (Table S6; Figure S4).

## Discussion

Longitudinal associations between social support and verbal memory were examined among English community-dwelling persons aged 50-89 years during 14-years follow-up. Our primary analyses produced four findings. First, in analyses of global social support, higher positive social support predicted higher baseline verbal memory and slower decline, whilst higher-than-usual levels of negative social support were modestly associated with faster decline. Secondly, verbal memory trajectories were gender and source-specific. Thirdly, differential rates of decline were primarily observed for differences between-persons in social support, not for within-person change. Fourthly, the direction of associations agreed with our hypotheses: higher positive and lower negative social support predicted higher baseline verbal memory and slower decline.

### Comparisons with previous studies

Recent meta-analyses confirmed that multiple aspects of poor social relationships (structural, functional, and combined measures) were associated with higher risk of cognitive decline (Kuiper *et al*., 2016). Heterogeneity in study populations, cognitive function domains, length of follow-up, and analytical techniques makes precise comparisons with previous studies difficult. Nevertheless, our findings agree to some extent with previous investigations.

With regards to global social support, our findings presented herein of (1) slower decline in verbal memory among participants with higher positive social support, and (2) better verbal memory performance at baseline among those with lower negative social support agree with our previous ELSA investigation (Liao and Scholes, 2017). The potential for emotionally supportive relationships to protect against cognitive decline has been established in analyses of ageing cohorts such as the MacArthur Studies of Successful Aging (Seeman *et al*., 2001) and the Longitudinal Aging Study Amsterdam (Ellwardt *et al*., 2013).

The gender- and source-specific associations presented here also agree with previous research suggesting that men and women perceive and utilise social support differently (Antonucci and Akiyama, 1987). Our findings of slower decline in verbal memory (1) among men with lower negative social support from a spouse/partner and from friends, and (2) among women with higher positive social support from extended family, match our previous investigation (Liao and Scholes, 2017). In agreement with our findings for men, analyses pooled over both genders showed that negative social support from close relationships predicted accelerated cognitive decline in the Whitehall II UK study (Liao *et al*., 2014), whilst positive social support from children (assessed at baseline) reduced risk of dementia over 10-years in the ELSA cohort (Khondoker *et al*., 2017). In the US HRS, being married/partnered and reporting more frequent contact with friends were independently associated with slower decline in episodic memory during 6-year follow-up (Zahodne *et al*., 2019).

### Potential mechanisms for the social support and cognitive ageing associations

Our reliance on observational data, including contemporaneous assessments of social support and verbal memory, means that we cannot ascribe causality to our findings. Nevertheless, several psychological, behavioural and physiological pathways have been suggested in the literature for the social relationships and cognitive ageing associations. First, stimulation of the brain, at least partially through the cognitive demands associated with social interactions, can impact positively on brain structure and cognitive reserve (Cotton et al., 2019) thereby lowering vulnerability and providing resilience to neuropathological damage (Stern, 2002). Secondly, social relationships may directly and positively impact on cognitive function through reinforcement of healthy lifestyle behaviours and increasing access to health information (‘main effect’ hypothesis) (Kuiper *et al*., 2016). Thirdly, the psychological and physiological ‘stress-buffering’ aspects of positive social relationships may impact positively on cognitive reserve through lowering levels of stress hormones such as cortisol (Kuiper *et al*., 2016). Whilst the ‘main effect’ hypothesis relates more to functional aspects of social relationships (e.g. benefits of having a large social network), the ‘stress-buffering’ hypothesis relates more to structural aspects (Kuiper *et al*., 2016).

In addition to these pathways, the roles of reverse causality (whereby cognitive decline weakens social support rather than being the cause) and residual confounding on our findings cannot be eliminated. We minimised the influence of reverse causality by excluding potentially cognitively unhealthy participants at baseline, doing so however limits generalizability of our findings to some extent (Zaninotto *et al*., 2018). With regards to residual confounding, we adjusted for a wide range of time-invariant variables, including SEP, healthy lifestyle behaviours (including alcohol), mobility limitations, and depressive symptomology.

### Methodological considerations in studying cognitive ageing

Several methodological considerations arise when using data from ageing cohorts to investigate correlates of cognitive decline. First, the LME models in our study show more favourable trajectories among those who completed more cognitive assessments. Whilst several previous investigations adjusted for practice effects (Alley, Suthers and Crimmins, 2007; Karlamangla *et al*., 2009), others did not (Liao *et al*., 2014; Zaninotto *et al*., 2018; Zheng *et al*., 2018). ‘True’ rates of cognitive decline may be underestimated if practice effects are not accounted for (Rabbitt *et al*., 2001). Comparisons between studies could be improved via adoption of standardised techniques for testing (Salthouse, 2015) and for adjustment (Vivot *et al*., 2016) to minimise the impact of practice effects on the associations of interest.

Secondly, as the follow-up period for ageing cohorts such as ELSA and HRS lengthens – resulting in a progressively more selectively healthy subset of participants – optimal use of the available data requires analytical techniques suited to including the sizeable and increasing proportions of cases with partial information due to item and unit nonresponse (Mostafa and Wiggins, 2015). In the present study we used MICE to handle item nonresponse and IPW-GEE (in sensitivity analyses) to compensate for attrition bias. Both methods provide robust and valid inference under the assumption that data are MAR. Although the LME and IPW-GEE estimates were similar in sign and magnitude in the present study for the social support terms (Figure S3), they are not directly comparable (Kurland *et al*., 2009; Rawlings *et al*., 2017). LME models (via the random effects) implicitly impute cognitive trajectories beyond death (i.e. for an immortal cohort); in contrast, GEE models estimate parameters in a dynamic (i.e. changing) cohort of survivors (Kurland *et al*., 2009). We recommend that analysts apply both approaches and compare estimates (Kurland *et al*., 2009).

Looking ahead to future research, possible extensions to our work include: (1) modelling approaches which combine IPW, MI, and GEE modelling to avoid restricting estimation to settings with monotone response patterns and no missing covariate data (Seaman *et al*., 2012; Han, 2016); (2) a structural equation modelling approach using full information maximum likelihood (FIML) estimation (Zaninotto *et al*., 2018); and (3) use of joint modelling where longitudinal outcomes are modelled jointly with a model for study dropout due to death (Rizopoulos, 2012; Raitanen *et al*., 2019).

### Implications for public health

Social support has been identified by the World Health Organization as a key social environmental factor that can enhance health, participation, and security as people age (WHO, 2002). It represents the main source of informal care for older adults to protect against progression of functional limitations (Hu & Li, 2020) and improve quality of life (Liao & Brunner, 2016). While the magnitude of the associations shown in this present study were modest, even minor differences in levels of cognitive functioning can over a period of several years substantially increase dependence and risk of adverse outcomes such as dementia (Edwards *et al*., 2005). Levels of social support and cognitive functioning are modifiable aspects of healthy ageing (Noguchi *et al*., 2019); therefore, it is imperative from a public health perspective to identify, design and implement interventions for older adults and their support network in order to maximise the protective role that supportive relationships can play in preventing cognitive decline. Boosting social support levels through digital engagement is one possible intervention area worthy of investigation (Liao et al., 2020).

### Strengths and limitations

Strengths of the present study include the benefits of analysing the ELSA cohort which includes its relatively large sample size, national representativeness, multiple and detailed measurements of social support available at each wave from different sources (allowing a more nuanced analysis than previous studies) and validated tests of verbal memory. Our analyses were strengthened by a longer follow-up period relative to other studies, allowing for robust measurement of within-person change in exposure and outcome; disentangling of within and between-person associations (Hoffman and Stawski, 2009); adjustment for practice effects which is recommended in the cognitive epidemiology literature (Rabbitt *et al*., 2001; Vivot *et al*., 2016); and (4) inclusion of sensitivity analyses to examine the impact of attrition bias.

Our findings however should be interpreted cautiously due to several limitations. First, the social support variables were based on self-reports. Our estimates could be subject to differential response bias if those with better verbal memory performance are more likely to positively bias their responses to the social support questions, thereby potentially upwardly biasing the magnitude of associations (Seeman *et al*., 2011). Secondly, adjustment for a wide range of confounders may have led to an underestimation of effect sizes since some of the impact of social support on verbal memory may be mediated through factors such as health behaviours and depressive symptomology. Thirdly, the large number of tests by fitting gender and source-specific models increased the probability of detecting false associations (Type 1 error). Fourthly, attrition is a limitation inherent to analyses of cognitive ageing. Whilst LME models are robust under the MAR assumption, our findings should still be interpreted in the context of an analytic sample that was increasingly selective over time, with those most healthy and affluent being most likely to remain. Fifthly, information on death via mortality linkage is currently available in the public datasets only up to wave 6; thereafter we could not identify those lost to the study through death. Finally, as in all observation studies, our findings could have been influenced by additional confounders such as personality characteristics that were not available.

## Conclusion

In conclusion, although modest in magnitude, our findings provide support for the notion that high levels of positive social support and low levels of negative social support can protect against slower decline in verbal memory. These findings can inform future research studies and intervention strategies designed to maximise the potential role of supportive relationships in achieving healthy cognitive ageing.

## Data Availability

ELSA datasets, including the harmonized dataset created as part of the gateway of global ageing data to facilitate cross-national comparisons, are available for free upon registration to the UK Data Service (https://www.ukdataservice.ac.uk/). Clemens, S., Phelps, A., Oldfield, Z., Blake, M., Oskala, A., Marmot, M., Rogers, N., Banks, J., Steptoe, A., Nazroo, J. (2019). English Longitudinal Study of Ageing: Waves 0-8, 1998-2017. [data collection]. 30th Edition. UK Data Service. SN: 5050. http://doi.org/10.5255/UKDA-SN-5050-17.

## Acknowledgements

The authors thank the interviewers and nurses, the participants in the English Longitudinal Study of Ageing, and colleagues at NatCen Social Research. We thank the original data creators, depositors, copyright holders, the funders of the Data Collections and the UK Data Archive for the use of data from English Longitudinal Study of Ageing. The original data creators, depositors or copyright holders bear no responsibility for the current analysis or interpretation.

## Consent and ethical approval

ELSA participants provided signed consent for taking part in the study and for linkage to mortality data, and ethical approval for each wave was granted by the London Multicentre Research Ethics Committee (MREC/01/2/91).

## Data sharing statement

ELSA datasets, including the harmonized dataset created as part of the gateway of global ageing data to facilitate cross-national comparisons, are available for free upon registration to the UK Data Service (https://www.ukdataservice.ac.uk/).

Clemens, S., Phelps, A., Oldfield, Z., Blake, M., Oskala, A., Marmot, M., Rogers, N., Banks, J., Steptoe, A., Nazroo, J. (2019). *English Longitudinal Study of Ageing: Waves 0-8, 1998-2017*. [data collection]. 30^th^ Edition. UK Data Service. SN: 5050. http://doi.org/10.5255/UKDA-SN-5050-17.

Analytical syntax to enable replication of the results using the datasets archived at the UK data service is available at https://www.medrxiv.org/XXX.

## Abbreviations

ELSA: English Longitudinal Study of Ageing
HRS: Health and Retirement study
GEE: generalized estimation equations
IPW: inverse probability weighting
PM: person-mean
WP: within-person

## Multiple imputation

Missing data on predictor variables (social support and confounders) was replaced using MICE (n=10 imputations). Imputation models used were: (1) predictive mean matching (PMM) algorithm (social support); (2) multinomial logistic regression (education and wealth); (3) logistic regression (healthy lifestyles; social participation and depression). Variables with complete data used to impute the missing values were as follows: gender; age; baseline verbal memory; mobility limitations; whether living as part of a couple; presence of any doctor-diagnosed health conditions; smoking status; and the wave 1 weight.

## Modelling

LME model on each imputed dataset estimated via maximum likelihood (weighted by wave 1 weight). Social support scores summed across all 4 sources (spouse/partner; children; friends; extended family). Estimates adjusted for time, time^2^, age, age^2^, (time × age; time × age^2^), number of prior word-recall assessments (practice effect), gender, education, wealth, smoking status, daily alcohol consumption, physical activity, social participation, depression and mobility limitations. Age at baseline was centred at age 65. The WP association is the estimated change in verbal memory on a measurement occasion when a participant’s level of social support is 1-unit higher than it usually is for them. The PM association is the estimated difference in verbal memory between persons who differ by 1-unit in their mean (‘stable’) level of social support. Estimates represent biennial change in verbal memory (i.e. change per every 2 years).

